# Neutralizing antibodies following three doses of BNT162b2 vaccine, breakthrough infection, and symptoms during the Omicron predominant wave

**DOI:** 10.1101/2022.09.15.22280009

**Authors:** Shohei Yamamoto, Kouki Matsuda, Kenji Maeda, Kumi Horii, Kaori Okudera, Yusuke Oshiro, Natsumi Inamura, Junko S. Takeuchi, Maki Konishi, Mitsuru Ozeki, Tetsuya Mizoue, Haruhito Sugiyama, Nobuyoshi Aoyanagi, Hiroaki Mitsuya, Wataru Sugiura, Norio Ohmagari

## Abstract

**Background:** Data on the role of immunogenicity following the third vaccine dose against Omicron infection and coronavirus disease 2019 (COVID-19)-compatible symptoms of infection are limited.

**Methods:** First we examined vaccine effectiveness (VE) of the third-dose against the second dose during the Omicron wave among the staff at a tertiary hospital in Tokyo. In a case-control study of a cohort of third vaccine recipients, we compared the pre-infection levels of live-virus neutralizing antibodies (NAb) against Omicron between breakthrough cases and their controls, who had close contact with COVID-19 patients. Among these cases, we examined the association between pre-infection NAb levels and the number of COVID-19-compatible symptoms experienced during the Omicron wave.

**Results:** Among the 1456 participants for VE analysis, 60 (4%) breakthrough infections occurred during the Omicron wave (January to March 2022). The third-dose VE for infection, relative to the second dose was 54.6% (95% CI: 14.0–76.0). Among the recipients of the third vaccine, pre-infection NAb levels against Omicron did not significantly differ between the cases and controls. Among the cases, those who experienced COVID-19-compatible symptoms had lower pre-infection NAb levels against Omicron than those who did not.

**Conclusions:** The third vaccine dose was effective in decreasing the risk of severe acute respiratory syndrome coronavirus 2 infection during the Omicron wave compared with the second dose. Among third-dose recipients, higher pre-infection NAb levels may not be associated with a lower risk of Omicron infection. Contrarily, they may be associated with fewer symptoms of infection.

**Summary:** The third vaccine dose reduced SARS-CoV-2 infection risk during the Omicron wave. Higher neutralizing antibody levels may not reduce Omicron infection risk in third-dose patients. On the contrary, it may be associated with fewer symptoms of infection.

## Introduction

During the ongoing coronavirus disease 2019 (COVID-19) pandemic, the waning of the second vaccine-induced immunogenicity over time [1, 2] and the emergence of variants of concerns (VOCs), such as Delta and Omicron, have led many countries to adopt the booster (third) vaccine campaign. Observational studies have shown that a third-dose of existing mRNA vaccines is still effective against infection with Delta and Omicron variants and hospitalization [3-5]; however, the third-dose vaccine effectiveness (VE) against Omicron infection is lower than that against Delta [3, 4]. The levels of vaccine-induced neutralizing antibodies (NAb) against Omicron were much lower than those against Delta and former severe acute respiratory syndrome coronavirus 2 (SARS-CoV-2) strains, even after the third-dose [6, 7], and such a low level of variant-specific immunogenicity may contribute to poor protection against Omicron infection.

Epidemiological evidence on the association between pre-infection humoral immunity and the risk of breakthrough infection during the Wuhan (wild-type) Alpha and Delta-dominant waves is inconsistent [8]. Several studies have shown an inverse relationship between vaccine-induced antibody levels and infection risk, whereas others have suggested that infection occurs irrespective of the antibody levels. Data regarding the association between vaccine-induced immunogenicity and infection risk during the Omicron epidemic are limited [9], which has increased transmissibility and high immune evasion. VE studies have reported a higher ratio of asymptomatic patients to overall infected patients in vaccinated COVID-19 patients than in unvaccinated patients [10, 11], suggesting a suppressive role of the vaccine against symptoms of infection. It is of interest to determine whether pre-infection NAb levels are a determinant of symptomatic episodes during breakthrough infection.

The aim of the present study was to investigate the role of immunogenicity following the third vaccine dose against Omicron infection and COVID-19-compatible symptoms. First, we examined VE of the third-dose relative to the second dose during the Omicron dominant wave among the staff of a tertiary hospital in Tokyo. In a case-control study nested in a cohort of third vaccine recipients, we compared the pre-infection live-virus Nab levels against Omicron following the third-dose between breakthrough cases and their controls who were in close contact with COVID-19 patients. In addition, we examined the association between pre-infection NAb and the number of COVID-19-compatible symptoms experienced during the Omicron wave.

## Methods

### Study setting

National Center for Global Health and Medicine, Japan (NCGM) is a medical research center for specific areas, including infectious diseases. As their mission, NCGM has played a major role in the care and research of COVID-19 since the early stage of the epidemic [12] and has accepted many patients with severe COVID-19. The in-house vaccination program using COVID-19 mRNA-LNP BNT162b2 (Pfizer-BioNTech) was conducted from March to June 2021 for the first and second doses and from December 2021 to February 2022 for the third vaccination.

In the NCGM, a repeat serological study was launched in July 2020 to monitor the spread of SARS-CoV-2 infection among staff during the COVID-19 pandemic. The details of the study have been reported elsewhere [13]. As of March 2022, we have completed five surveys, each of which measured the levels of anti-SARS-CoV-2 nucleocapsid- and spike- (from the second survey onward) protein antibodies for all participants using both the Abbott and Roche assays, the serum samples were stored at −80□, and information on COVID-19-related factors (vaccination, occupational infection risk, infection prevention practices, history of close contact with COVID-19 patients, COVID-19-compatible symptoms, and so on) were collected using questionnaires. Self-reported vaccination status was validated using the information provided by the NCGM Labor Office, if any. We identified COVID-19 cases among study participants from COVID-19 patient records maintained by the NCGM Hospital Infection Prevention and Control Unit, which provided information on the date of diagnosis, diagnostic procedure, possible route of infection (close contact), symptoms, hospitalization, return to work for all cases, and virus strain and cycle threshold (Ct) values for those diagnosed at the NCGM. Written informed consent was obtained from all participants. This study was approved by the NCGM ethics committee (approval number: NCGM-G-003598).

### Study for the effectiveness of the third-dose vaccine relative to the second dose

To examine the VE of the third versus second vaccine doses during the Omicron BA.1 dominant wave, we analyzed the data of NCGM staff who participated in the fifth survey (March 2022), for whom the detailed information on vaccine status was available. The outcome was polymerase chain reaction (PCR)- or antigen-confirmed COVID-19, which was reported in the in-house registry. In Japan, the Omicron dominant wave began in January 2022. Thus, we started the follow-up on January 1, 2022 (baseline) and censored the date of COVID-19 diagnosis or the attendance date to the fifth survey (March 1–9, 2022), whichever came first.

Of the 1578 participants, we excluded those who received less than two vaccine doses (n=30) at baseline (i.e., January 1, 2022), received non-mRNA-based COVID-19 vaccines (n=49), had a history of COVID-19 before the start of the follow-up (n=21), and tested positive on anti-SARS-CoV-2 nucleocapsid protein assays (positive with Abbott and/or Roche) in any previous survey (i.e., first to fourth surveys) (n=41), leaving 1456, who were included in the analysis.

### A nested case-control study among third-dose vaccine recipients

We conducted a case-control study among the staff who participated in the fourth survey in December 2021, after receiving the third dose (**Figure 1**). Of the 948 participants, 22 received the third vaccine of BNT162b2 at least seven days before the fourth survey and donated a blood sample. Among these participants, those with a history of COVID-19 (n = 3) and those who tested positive with the anti-SARS-CoV-2 nucleocapsid protein antibody assay (Abbott and/or Roche) in any previous survey (n = 3) were excluded. The remaining 218 infection-naïve participants formed the basis of this case-control study and attended the fifth survey (follow-up) conducted in March 2022.

**Figure 1.**
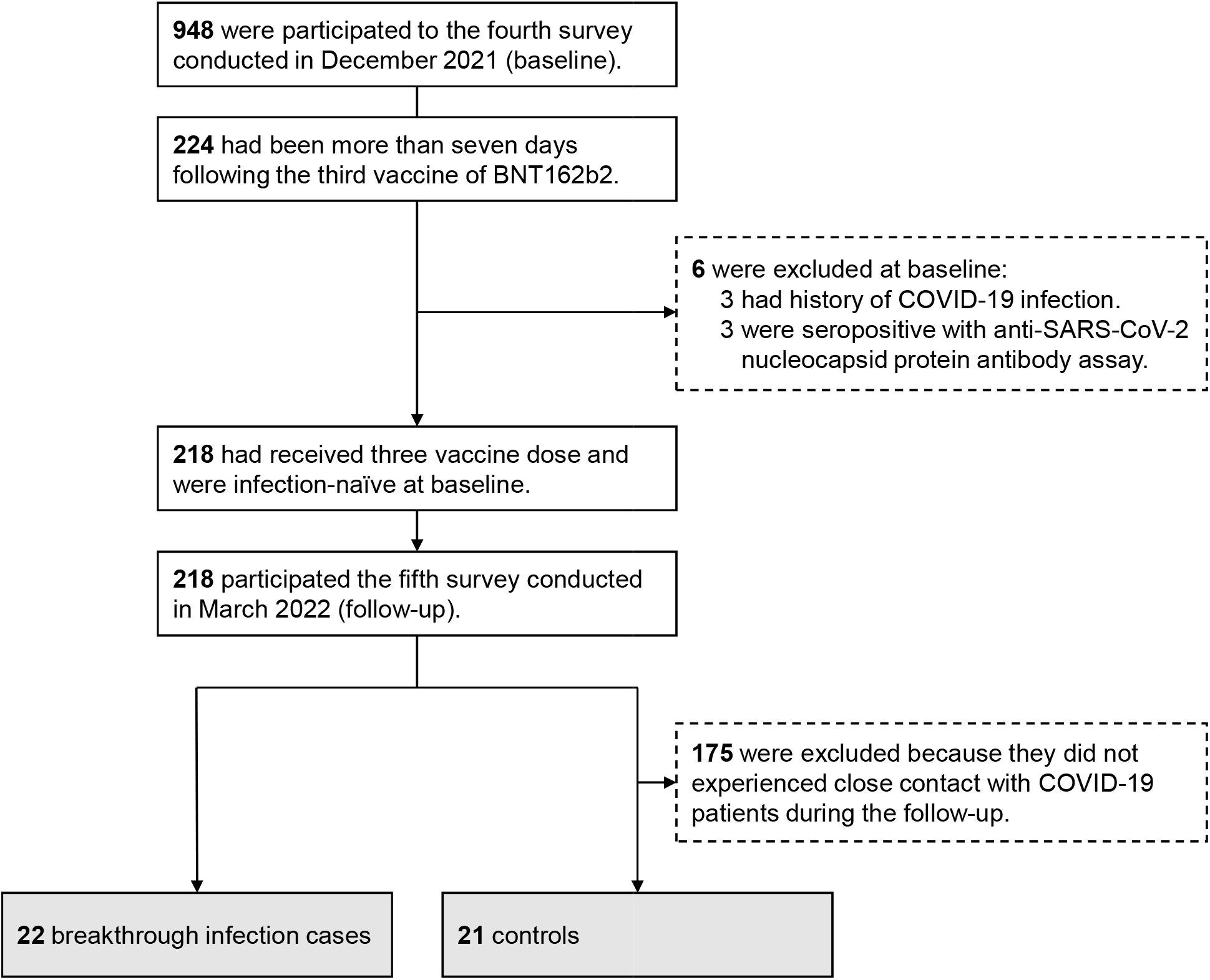
Flowchart for the case-control selection

In this cohort, 22 breakthrough infections were identified during the follow-up. Of these, 10 cases were ascertained via an in-house registry: nine were tested positive by PCR testing and one by antigen test. The remaining 12 patients were identified using anti-SARS-CoV-2 nucleocapsid protein assays (Abbott and/or Roche) during the follow-up survey. Five out of ten cases confirmed via the in-house registry were sequenced and confirmed to be infected with the Omicron BA.1 variant. We could reasonably assume that nearly all cases were infected with Omicron variants, which accounted for more than 90% of the sequenced COVID-19 samples in Japan during the study period [14]. Of the remaining 196 participants without evidence of COVID-19, 21 participants reported that they had close contact with COVID-19 patients during the follow-up and served as controls.

NAb in post-third vaccine and pre-infection serum was determined by quantifying serum-mediated suppression of the cytopathic effect (CPE) of each SARS-CoV-2 strain in VeroE6TMPRSS2 cells [15, 16]. The routes of the cells and viruses are described in Supplemental Text 1. Each serum sample was 4-fold serially diluted in a culture medium. The diluted sera were incubated with 50% tissue culture infectious dose (TCID_50_) of the virus at 37 °C for 20 minutes (final serum dilution range of 1:40–1:25000), after which the serum-virus mixtures were inoculated into VeroE6_TMPRSS2_ cells (1.0×104/well) in 96-well plates. The SARS-CoV-2 strains used in the assays were as follows: Wuhan, wild-type strain (SARS-CoV-2^05-2N^) [17], Omicron BA.1 variant (SARS-CoV-2^TKYX00012/2021^), and Omicron BA.2 variant (SARS-CoV-2^TKYS02037/2022^). After cell culture for 3–5 days, the CPE levels observed in cells exposed to SARS-CoV-2 were determined using the WST-8 assay with a Cell Counting Kit-8 (Dojindo, Kumamoto, Japan). The serum dilution that resulted in 50% inhibition of CPE was defined as the 50% neutralization titer (NT_50_). Each serum sample was tested in duplicate, and the average value was used for the analysis.

We assessed anti–SARS-CoV-2 antibodies for all participants at the baseline and follow-up and retrieved the data for the case-control subsets. We quantitatively measured the levels of antibodies against the receptor-binding domain of the SARS-CoV-2 spike protein using the AdviseDx SARS-CoV-2 IgG II assay (Abbott) (immunoglobulin [Ig] G [IgG]) and Elecsys® Anti-SARS-CoV-2 S RUO (Roche) (including IgG).

We also qualitatively measured antibodies against the SARS-CoV-2 nucleocapsid protein using the SARS-CoV-2 IgG assay (Abbott) and Elecsys^®^ Anti-SARS-CoV-2 RUO (Roche), used these data to exclude those with possible infection before the baseline survey and to identify those who experienced breakthrough infection during the follow-up period. The sensitivity and specificity were 100% and 99.9%, respectively, for the Abbott assay [18] and 99.5% and 99.8%, respectively, for the Roche assay [19].

### Symptoms indicative of COVID-19

To detect COVID-19-compatible symptoms among breakthrough infection cases, we collected information on symptoms using a questionnaire during the follow-up survey. In the questionnaire, we asked participants about the following symptoms since January 1, 2022: (1) common cold-like symptoms (sore throat, cough, runny nose, nasal congestion, fever, and fatigue.) persisting for four or more days, (2) common cold-like symptoms persisting for less than four days, (3) high fever, (4) severe fatigue, (5) dyspnea, (6) loss of sense of taste or smell, and (7) sore throat. In the questionnaire, we highlighted that adverse reactions after vaccination should not be included. For the analysis, responses to two questions regarding common cold-like symptoms were combined and categorized into three groups: no symptoms, persists for <4 days, and persists for ≥4 days. We counted five specific symptoms: high fever, severe fatigue, dyspnea, taste and smell disorders, and sore throat and classified them into three categories: 0, 1–2, and 3–5. We did not inquire about the timing of the symptoms onset during the follow-up. For cases notified via the in-hospital registry, we confirmed that the self-reported symptoms in the follow-up survey were similar to those at COVID-19 diagnosis recorded in the registry.

### Statistical analysis

In the study on the VE of the third vaccine dose relative to the second dose, we fitted a Cox proportional hazards model with adjustment for age and sex. We treated vaccination status as a time-dependent covariate. If the participants received the third vaccine dose during the follow-up period, the vaccination status was changed from two to three doses 14 days after the date of receiving the third-dose. The estimated hazard ratio (HR) with a 95% confidence interval (CI) for vaccination status was used to calculate VE (%) according to the following formula: VE = (1 – adjusted HR) × 100.

In the nested case-control study among third-dose vaccine recipients, we compared the characteristics between cases and controls using the Mann–Whitney U test or Fisher’s exact test. To examine the difference in the humoral response after the third vaccination between cases and controls, we compared the log-transformed titers of live-virus NAb (Wuhan, Omicron BA.1, and Omicron BA.2) and anti-spike antibodies (Abbott and Roche) using a multivariable linear regression model with adjustment for age (continuous) and sex (male or female). We also constructed a multivariate linear regression model to examine the association between pre-infection Nab titers after the third vaccination and COVID-19-compatible symptoms during the study period among the cases. To compare the inter-individual differences in Nab titers against Wuhan, Omicron BA.1, and Omicron BA.2, between cases and controls, we used a generalized estimating equation (GEE) with a robust variance estimator. The estimated effects of covariates were back-transformed and presented as ratios of geometric means and geometric mean titers (GMTs) with 95% CIs. For sensitivity analysis, we compared pre-infection NAb and anti-spike antibody titers between cases and controls, restricting those who experienced close contact at home, where virus transmission would be higher than that in other settings.

For analyses, values below or above the limit of detection (LOD) for NAb titers (NT_50_<40) and spike antibody titers with the Roche assay (titer >25000 U/mL) were given the LOD value. Statistical analyses were performed with Stata version 17.0 (StataCorp LLC), and graphics were prepared with GraphPad Prism 9 (GraphPad, Inc.). All *P*-values were two-sided, and statistical significance was set at *P* < 0.05.

## Results

### Effectiveness of the third dose relative to the second dose

Among the 1456 infection-naïve participants who received two or three vaccine doses, 60 (4%) SARS-CoV-2 breakthrough infections occurred between January 1 and March 9, 2022 (**Table 1 and Figure S1**). Regarding the variant types, 33 (55%) were identified as Omicron, and 27 were unknown (unmeasured). The median age of the patients was 39 years (interquartile range [IQR], 28–45 years), and 80% were females. At the beginning of the follow-up, the median intervals since the last vaccination in the two and three dose groups were 261 (IQR: 233–267) and 15 (IQR: 15–19) days, respectively. Fourteen recipients who received two doses were infected at a median of 218 (IQR: 160–255) days after the second vaccination, whereas 46 cases among the three-dose vaccine recipients were infected at a median of 55 (IQR: 43–61) days after the third vaccination. Regarding the symptoms during infection identified from the in-house registry, all patients were asymptomatic (n=10, 17%) or had mild symptoms (n=50, 83%). Three patients with mild symptoms (6 %) were admitted to a hospital. The proportion of asymptomatic patients of overall infected patients was higher in patients who received three doses (20%) than in those who received two doses (7%). The infection rates per 10,000 person-days for the two and three vaccine doses were 9.7 and 6.3, respectively, and the age- and sex-adjusted VE of the three doses against two doses for infection was 54.6% (95% CI: 14.0–76.0).

**Table 1.** Vaccine effectiveness of three versus second doses of vaccine for COVID-19 infection among 1456 staff of a large referral medical and research institution in Tokyo during the Omicron wave (January 2022 to March 2022).

| Vaccination status | Total |  | COVID-19 cases |  |  | Person-days | Incident rate per 10000 person-days | Adjusted VE, % (95% CI) |
| --- | --- | --- | --- | --- | --- | --- | --- | --- |
|  | No. | Interval from last vaccination to tracking start date, median days (IQR) | No. | Interval from last vaccination to infection, median days (IQR) | Asymptomatic cases, n (%) |  |  |  |
| Two doses | 863 | 261 (233–267) | 14 | 218 (160–255) | 1 (7) | 14429 | 9.7 | reference |
| Three doses | 1375 | 15 (15–19) | 46 | 55 (43–61) | 9 (20) | 73645 | 6.3 | **54.6 (14.0–76.0)** |

Vaccine effectiveness (VE) was calculated as $([1 - \text{hazard ratio}] \times 100\%)$ , estimated using a time-dependent Cox proportional hazard regression model with adjustment for age and sex.
In the two dose group (n=863), 782 received the third-dose during the follow-up without infection and moved to the three dose group.

### Characteristics of breakthrough infection in the case-control analysis

Of the 22 patients with breakthrough infections after the third vaccine dose, 86% were male with a median age of 31.8 (IQR: 26.6–38.4) years, all were nurses, and 23% were affiliated with the COVID-19-related department (**Table 2**). The majority (91–100%) showed good adherence to infection-prevention practices. In the follow-up survey, 36% reported having spent 30 min or more in crowded places, close-contact settings, and confined and enclosed spaces (the 3Cs) without a mask, whereas 27% reported having dined in a group of five or more people for more than one hour. In the follow-up survey, 23% of respondents reported close contact with COVID-19 patients in their households, 5% in the community, and 5% in the workplace. These figures were similar between cases and controls, except for those with close contact; close contact in the workplace setting was more frequent in controls than in cases (57% vs. 5%).

**Table 2.**
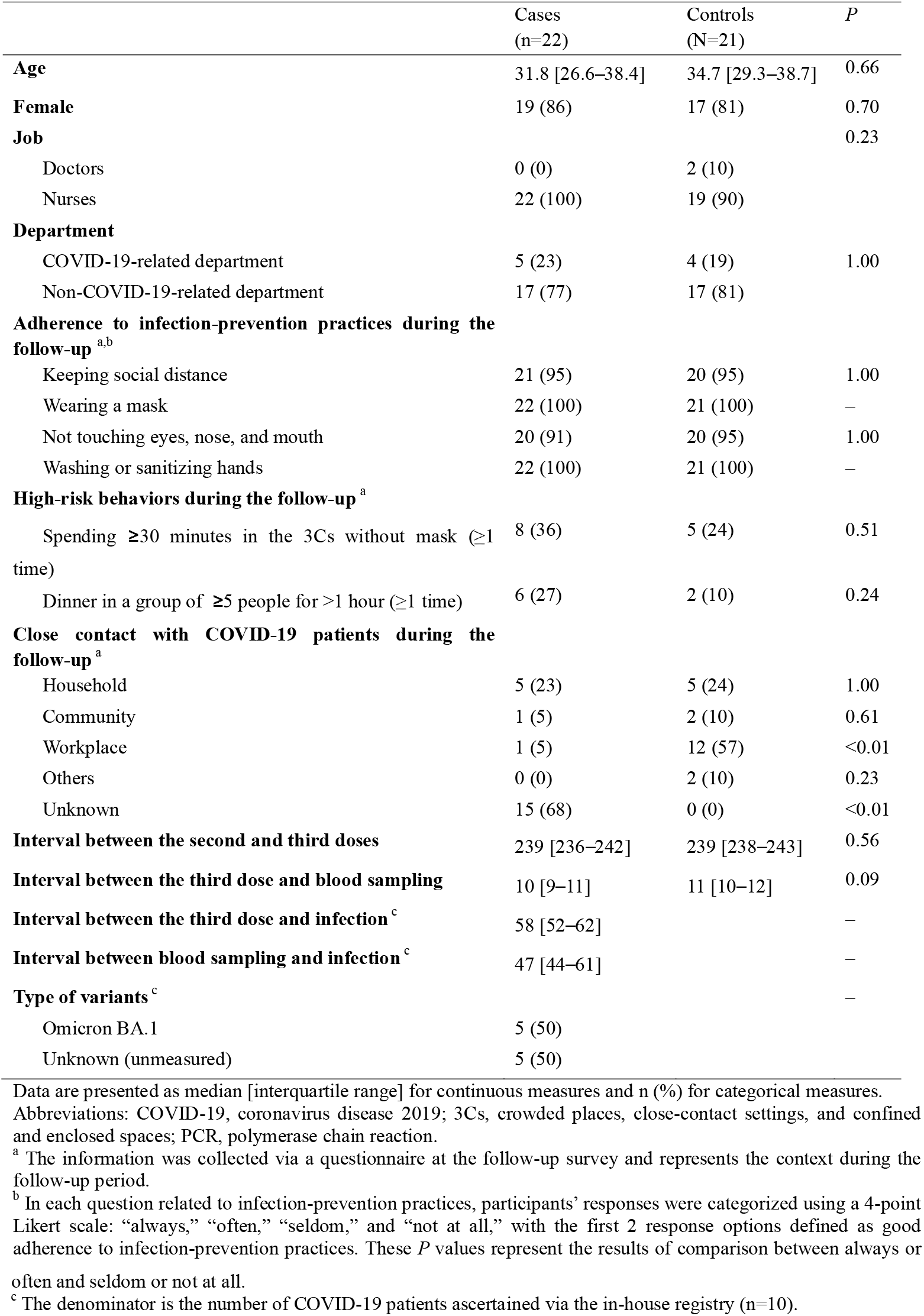
Characteristics of the participants in the case-control study

As for 10 cases that were notified from the in-house COVID-19 registry, the median intervals from the third vaccine to the infection and the blood sampling to the infection were 58 (IQR: 52–62) and 47 (IQR: 44–61) days, respectively. Of these, 10% were asymptomatic, and 90% had mild symptoms. None of the patients required hospital admissions. Of the 12 cases identified solely by antibody test, 42% reported some COVID-19-compatible symptoms during follow-up: sore throat (n=5), common cold-like symptoms (n=4), severe fatigue (n=2), and dyspnea (n=1).

### Neutralizing and anti-spike antibodies in cases and controls

There were no statistical differences between the cases and controls in the pre-infection levels of NAb against Wuhan, Omicron BA.1, or Omicron BA.2 or anti-spike antibodies (**Figure 2 and Supplementary Table S1**). The GMT (NT_50_) of NAb against Wuhan, predicted by the linear regression with adjustment for age and sex, was 733 (95% CI, 552–973) for cases and 613 (95% CI, 407–923) for controls, and the case-to-control ratio was 1.20 (95% CI, 0.72–1.98). That against Omicron BA.1 was 221 (95% CI, 141–349) for cases and 223 (95% CI, 129–383) for controls, with a ratio of 1.00 (95% CI, 0.49–2.02). That against Omicron BA.2was 102 (95% CI, 72–143) for cases and 108 (95% CI, 75–155) for controls, and the ratio was 0.94 (95% CI, 0.57–1.55). The GMT of the anti-spike antibody with the Abbott assay (arbitrary units [AU]/mL) was 23290 (95% CI, 19412–27942) for cases and 22670 (95% CI, 17411–29519) for controls, with a ratio of 1.03 (95% CI, 0.74–1.42). The GMT with the Roche assay (U/mL) was 21143 (95% CI, 18821–23752) for cases and 19692 (95% CI, 16660–23276) for controls, and the ratio was 1.07 (95% CI, 0.87–1.32). The results were the same among individuals who had close contact with family members with COVID-19 (**Supplementary Table S2**).

**Figure 2.**
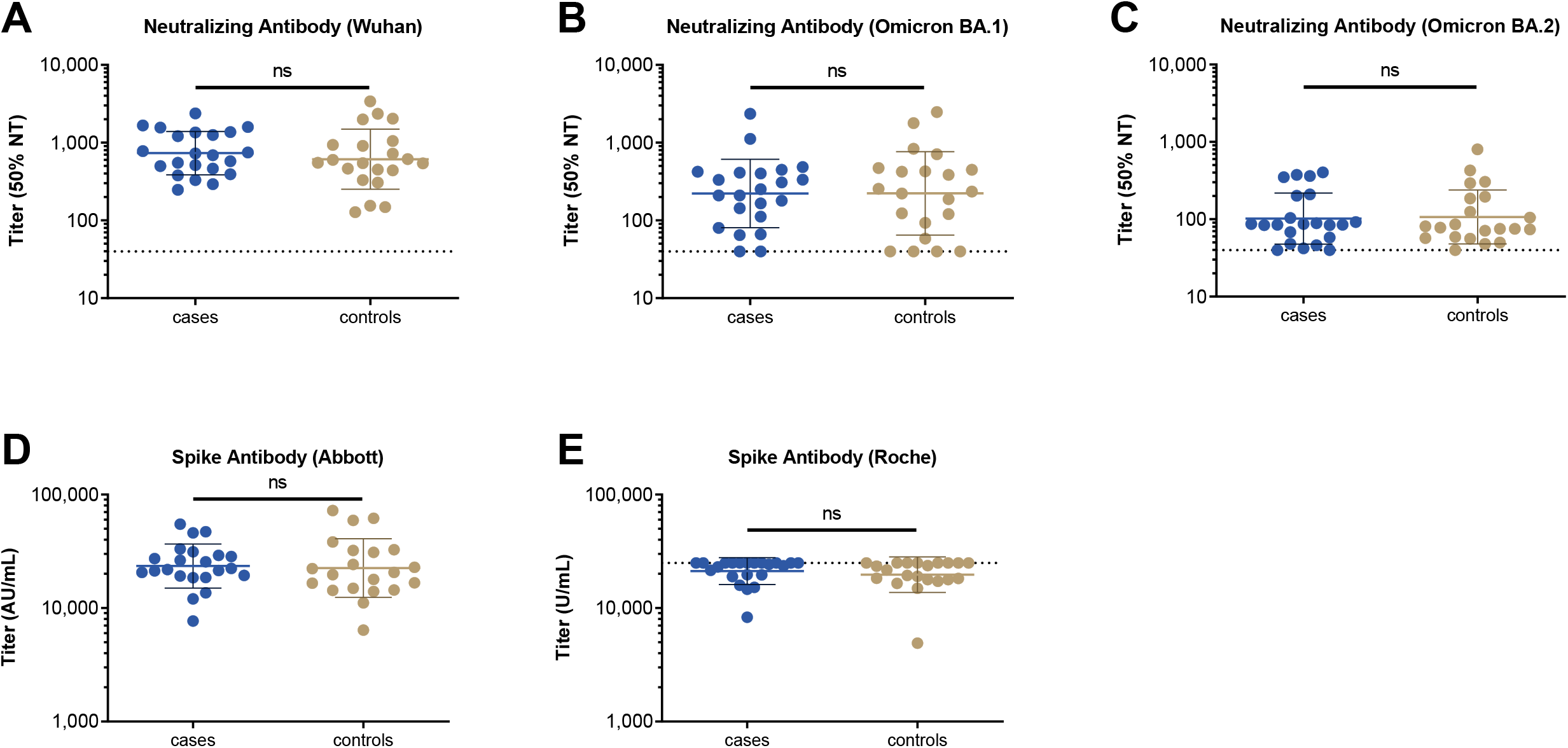
Pre-infection neutralizing and spike antibody titers following the third vaccine dose among cases and controls who experienced close contact with COVID-19 patients. Among the 22 SARS-CoV-2 breakthrough infection cases and the 21 controls who experienced close contact with COVID-19 patients, neutralizing antibody titers against the Wuhan (A), Omicron BA.1 (B), and Omicron BA.2 (C) strains following the third vaccine dose during the pre-infection period (median of 10 days since the third vaccination) are shown. Results of comparison of post-vaccination anti-spike antibody titers measured using the Abbott reagent (D) and those using the Roche reagent (E) in the two groups. In each panel, the horizontal bars indicate the geometric mean titers, and the I-shaped bars indicate geometric standard deviations. The dashed horizontal lines indicate the limits of detection in the present analysis (NT_50_<40 in neutralizing antibody assays and >25000 U/mL in Roche assay). *P* values were calculated using a multivariable linear regression model with adjustment for age and sex (ns: not significant). Abbreviations: AU, arbitrary units; COVID-19, coronavirus disease 2019; NT, neutralizing titer; SARS-CoV-2, severe acute respiratory syndrome coronavirus 2.

### Cross-reactive neutralizing antibodies

NAb titers against Omicron BA.1 and Omicron BA.2 were much lower than those against Wuhan (**Figure 3)**. Among the cases, the NAb titers against Omicron BA.1 and Omicron BA.2 were 3.3-fold and 7.2-fold lower than those against Wuhan, respectively. Those against Omicron BA.2 were 2.2-fold lower than those against Omicron BA.1. Similar results were obtained in the control group.

**Figure 3.**
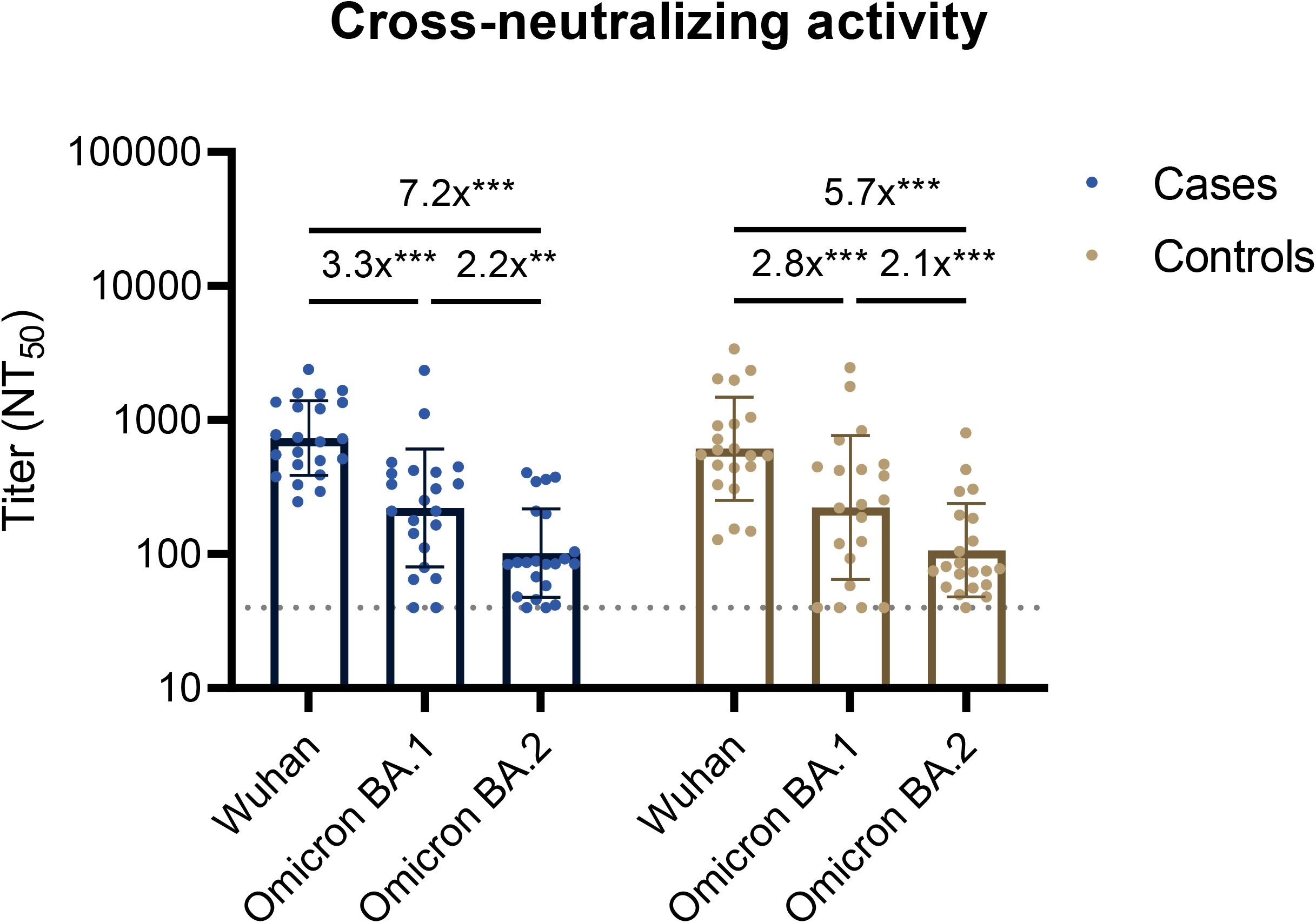
Cross-reactive neutralizing antibody titers following the third vaccine dose among cases (n=22) or controls (n=21). Results of comparison of pre-infection neutralizing antibody titers against the Wuhan, Omicron BA.1, and Omicron BA.2 strains among cases or controls. The bars indicate geometric mean titers, and I-shaped bars indicate their geometric standard deviations. The dashed horizontal lines indicate the limits of detection (NT_50_ titer <40). The fold-change values are estimated ratios of geometric means for antibody titers based on the GEE model (ns: not significant; *P<0.05; **P<0.01; ***P<0.001). Abbreviations: GEE, generalized estimating equation; NT_50_, 50% neutralization titer; m, months

### Neutralizing antibodies and symptoms in patients in the case-control study

Among cases identified in the case-control analysis, those who experienced some COVID-19 compatible symptoms during the study period had lower, albeit not statistically significant, pre-infection NAb levels against the Omicron BA.1 and BA.2 than those who did not experience any symptoms (**Table 3**). Those who experienced common cold-like symptoms had significantly lower pre-infection NAb titers against Omicron BA.1 than those who did not have these symptoms. Compared to those who did not experience common cold-like symptoms, the GMT (95% CIs) ratio of NAb against the Omicron BA.1 for those who had the symptoms for less than four days and for four or more days was 0.81 (0.04–15.1) and 0.39 (0.20–0.77), respectively (*P* for trend <0.01). Additionally, a larger number of symptoms were associated with lower pre-infection NAb titers against Omicron BA.1. Although not significant, similar associations of common cold-like symptoms and the number of symptoms were observed with NAb titers against Omicron BA.2. High fever was significantly associated with lower NAb titers against Omicron BA.2. The loss of taste or smell was associated with lower Omicron BA.1 titers.

**Table 3.**
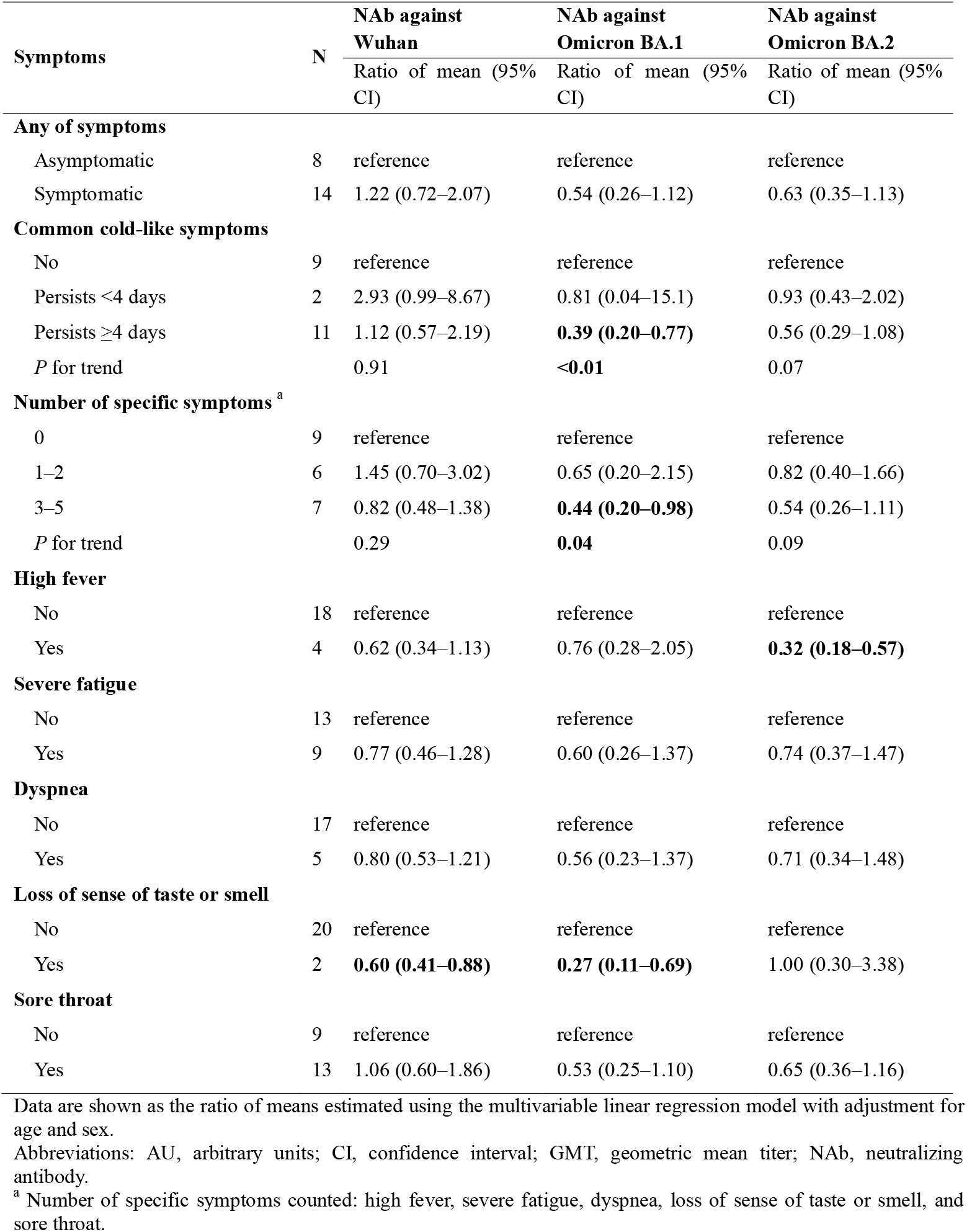
Association between pre-infection neutralizing antibody titers following the third vaccine dose and symptoms among breakthrough infection cases.

## Discussion

Among the staff of a medical research center in Tokyo, the incidence rate of breakthrough infection with SARS-CoV-2 during the Omicron dominant wave was lower in those who received three doses of the mRNA vaccine (within 3 months following vaccination) than in those who received only two doses (within 11 months following vaccination). In a case-control study nested in a cohort of vaccine recipients of all three doses, the pre-infection NAb levels against Omicron BA.1 and Omicron BA.2 did not materially differ between breakthrough infection cases and uninfected controls who were in close contact with COVID-19 patients during the Omicron wave. The patients who experienced COVID-19-compatible symptoms during the Omicron wave had a lower pre-infection NAb levels against Omicron than those without symptoms.

During the Omicron BA.1 dominant wave, we observed 54.6% VE of the third dose of vaccine (within three months of vaccination) compared with the second dose (within 11 months of vaccination). This finding is similar to those of previous studies that examined VE approximately one–two months after the third-dose relative to unvaccinated individuals during the Omicron wave (51.2 and 52.9%) [3, 4]. Our results were consistent with our laboratory data from the same cohort [20], where Omicron BA.1-specific NAb was detected in all patients who received the third-dose within one month but not in all patients who received only the second dose approximately eight months prior (GMT, 152 v.s. <40 NT_50_). However, compared with the VE during the Delta epidemic (88.1 and 92.6%) [3, 4], VE during the Omicron epidemic was considerably low. In serum samples obtained from the third-dose vaccine recipients, we found much lower NAb titers against Omicron BA.1 than against Delta (152 v.s. 1563 NT_50_) [20]. Thus, the third vaccine dose was effective in decreasing the risk of Omicron infection. However, the magnitude of this effect was much smaller than that for former SARS-CoV-2 strains.

Among the recipients of the third-dose who had close contact with COVID-19 patients during the Omicron wave, we observed no measurable differences in the levels of live-virus Nab against Omicron variants and anti-spike antibodies between breakthrough infection cases and their controls. This result is consistent with the findings of a study of nursing home residents, which showed no difference in pseudo typed virus NAb titers against Omicron following the third vaccination between Omicron-infected patients and arbitrarily selected controls [9]. These results suggest that among healthy people, breakthrough infections with highly immune-evasive Omicron variants may occur independent of the humoral immune response to the third-dose vaccine.

In our VE analysis, asymptomatic infection was more frequent among recipients of the third-dose (20%) than among those of the second dose (7%), which is consistent with previous data [21, 22]. Among the cases in our case-control study of third vaccine recipients, higher pre-infection NAb titers against Omicron was associated with fewer symptoms during the Omicron wave. Similarly, in a study of patients infected with Omicron after vaccination, a higher total antibody level within seven days of infection (i.e., peri-infection period) was associated with a lower rate of fever, hypoxia, CRP elevation, and lymphopenia [23]. These findings suggest that pre-infection NAb may play a role in alleviating symptoms of SARS-CoV-2 infection.

The present study has several strengths. We measured NAb titers against Wuhan, Omicron BA.1, and Omicron BA.2 using live viruses. Both cases and controls were derived from a well-designed cohort. The cases were identified through the COVID-19 patient registry and a serological survey. This point is important because many patients with Omicron infection, characterized by asymptomatic or mild symptoms [24], might have been left undiagnosed. Additionally, we excluded undiagnosed cases from controls using the results of the antibody test at the follow-up.

This study has some limitations. First, we measured antibody levels at a median of 10 days following the third dose; antibody titers may still be rising, and we did not measure peri-infection antibody levels. Second, although we selected controls from among those who had close contact with COVID-19 patients, the major setting of the contact differed between cases (unknown) and controls (workplace). Nonetheless, we confirmed no material change in the results after restricting the cases and controls to those who had close contact at home (**Supplementary Table S2**). Third, among the cases reported from the in-house registry, viral sequence data were available for only 33 (55%) cases in the VE analysis and 5 (50%) cases in the case-control analysis. Nevertheless, we could reasonably assume that the remaining breakthrough infections were also due to the Omicron variant, which accounted for more than 90% of the sequenced COVID-19 samples in Japan during the follow-up period (January–March 2022) [14]. Fourth, we did not assess the cellular immune response, which is another important mechanism for preventing COVID-19 [25]. Finally, we could not determine whether the COVID-19-compatible symptoms reported in the follow-up survey corresponded to those that occurred during infection.

In conclusion, compared with that in the second dose, the third vaccine dose halved the risk of SARS-CoV-2 infection during the Omicron wave. Among the third-dose recipients, pre-infection and live-virus Nab titers against Omicron were not materially different between the cases and controls, whereas a higher pre-infection NAb titer against Omicron was associated with fewer symptoms. Higher levels of NAb after the third vaccination may not indicate a lower risk of Omicron infection, whereas they may suppress symptomatic episodes of infection.

## Data Availability

All data produced in the present study are available upon reasonable request to the authors.

## Acknowledgements

We thank Mika Shichishima, Yumiko Kito, and Azusa Kamikawa for their contribution to data collection and the staff of the Laboratory Testing Department for their contribution to measuring antibody testing.

